# Improved Sensitivity, Safety, and Rapidity of COVID-19 Tests by Replacing Viral Storage Solution with Lysis Buffer

**DOI:** 10.1101/2020.09.25.20201921

**Authors:** Oran Erster, Omer Shkedi, Gil Benedek, Eyal Zilber, Itay Varkovitzky, Rachel Shirazi, Dorit Oriya Shorka, Yuval Cohen, Tzahi Bar, Rafi Yechieli, Michal Tepperberg Oikawa, Dana Venkert, Michal Linial, Esther Oiknine-Djian, Michal Mandelboim, Zvi Livneh, Gilat Shenhav-Saltzman, Ella Mendelson, Dana Wolf, Moran Szwarcwort-Cohen, Orna Mor, Yair Lewis, Danny Zeevi

**Affiliations:** Central Virology Laboratory, Ministry of Health, Chaim Sheba Medical Center, Tel-Hashomer, P.O. Box 5265601, Ramat-Gan, Israel; Faculty of Medicine, Technion - Israel Institute of Technology, Haifa, Israel; Hadassah Medical Center, Jerusalem, Israel; Department of Internal Medicine, Sheba Medical Center, Tel Hashomer, Israel; Directorate of Defense Research & Development, Israeli Ministry of Defense, Israel; Independent Researcher, Israel; Department of Neurobiology, Faculty of Life Sciences, Tel Aviv University, Tel Aviv 69978, Israel; Department of Biological Chemistry, Institute of Life Sciences, The Hebrew University of Jerusalem, Jerusalem, Israel; School of Public Health, Sackler Faculty of Medicine, Tel-Aviv University, Tel-Aviv, Israel; Department of Biomolecular Sciences, Weizmann Institute of Science, Rehovot 7610001, Israel; Sackler Faculty of Medicine, Tel Aviv University, Tel Aviv, Israel; Virology Laboratory, Rambam Health Care Campus, Haifa, Israel; Navina AI Medical Technologies, Tel Aviv, Israel; Department of Biotechnology, Hadassah Academic College, Jerusalem, Israel

## Abstract

Conducting numerous, rapid, and reliable PCR tests for SARS-CoV-2 is essential for our ability to monitor and control the current COVID-19 pandemic.

Here, we tested the sensitivity and efficiency of SARS-CoV-2 detection in clinical samples collected directly into a mix of lysis buffer and RNA preservative, thus inactivating the virus immediately after sampling.

We tested 79 COVID-19 patients and 20 healthy controls. We collected two samples (nasopharyngeal swabs) from each participant: one swab was inserted into a test tube with Viral Transport Medium (VTM), following the standard guideline used as the recommended method for sample collection; the other swab was inserted into a lysis buffer supplemented with nucleic acid stabilization mix (coined NSLB).

We found that RT-qPCR tests of patients were significantly more sensitive with NSLB sampling, reaching detection threshold 2.1±0.6 (Mean±SE) PCR cycles earlier then VTM samples from the same patient. We show that this improvement is most likely since NSLB samples are not diluted in lysis buffer before RNA extraction. Re-extracting RNA from NSLB samples after 72 hours at room temperature did not affect the sensitivity of detection, demonstrating that NSLB allows for long periods of sample preservation without special cooling equipment. We also show that swirling the swab in NSLB and discarding it did not reduce sensitivity compared to retaining the swab in the tube, thus allowing improved automation of COVID-19 tests. Overall, we show that using NSLB instead of VTM can improve the sensitivity, safety, and rapidity of COVID-19 tests at a time most needed.

## Introduction

Rapid and robust identification of individuals infected by COVID-19 is one of the mainstays of containment and mitigation efforts of this pandemic.

The current guidelines of the CDC and WHO for SARS-CoV-2 tests are that collected swabs should be placed in a transport tube that is either empty or contains either Viral Transport Medium (VTM), Amies transport medium, or sterile saline [1,2]. Under such conditions, the viral capsid remains intact and active. While this strategy is the mainstay of clinical testing protocols for many pathogens, as it allows culturing of the pathogens of interest, there are disadvantages to be considered. Since the virus is kept in its infectious state, the specimens pose a significant biohazard both during transport and in the lab and require special safety procedures of packaging, transport, and treatment under BSL2 conditions in the lab which cause a considerable bottleneck in the processing workflow. After unpacking in the lab, a small fraction of the transport medium with the specimen is transferred into Lysis Buffer (LB) which inactivates the virus but also dilutes the sample typically ∼2-3 fold. This leads to a smaller overall quantity of viral RNA used in the diagnostic test, potentially leading to a higher proportion of false-negative results, especially in borderline cases. In addition, transport in VTM or any of the other common transport mediums requires the specimen to be kept at 4°C in order to prevent degradation of viral nucleic acid. This can present logistic challenges in the testing process.

We hypothesized that the collection of SARS-CoV-2 clinical specimens into a tube containing a Nucleic Acid Stabilization and Lysis Buffer (NSLB), instead of VTM, will streamline the clinical diagnostic workflow as this buffer both inactivates the virus and preserves viral RNA at room temperature. The use of NSLB will allow transport of the specimens without the need for cooling and improve the biosafety profile of the entire diagnostic workflow. Additionally, this buffer obviates the need for incubation in LB and the samples can be directly inserted into the RNA extraction step. This leads to a larger quantity of viral nucleic acid per sample in the diagnostic pathway.

Here, we compared the performance of RT-qPCR on samples collected into VTM and NSLB from 77 COVID-19 patients and 20 healthy controls and observed a significantly higher sensitivity for NSLB over VTM.

## Methods

In order to compare sampling into VTM vs. NSLB, we sampled each participant consecutively twice and put one specimen into VTM and one into NSLB (in alternating order). Each specimen was obtained from nasal and oropharyngeal swabs combined into one tube. We tested alternately two different commercially available NSLBs (DNA/RNA shield from ZYMO, and PrimeStore MTM from LongHorn).

Participants were recruited from three medical centers in Israel. The participants included 26 hospitalized patients with moderate to severe symptoms from the Sheba Medical Center, 21 hospitalized patients from Hadassah Medical Center, and 32 asymptomatic or mildly symptomatic patients who were quarantined in a hotel (their samples were processed by the Rambam Medical Center). All patients were found positive to SARS-CoV-2 by RT-qPCR prior to participating in this study. In addition, we recruited 20 healthy volunteers. All participants supplied written consent and the experiments were approved by the IRBs of the Sheba, Hadassah, and Rambam Medical Centers. Sampling and RT-qPCR for each of the three experiments were conducted by different teams at different labs.

Samples were collected 2-8 hours before RNA extraction in the lab. During that time, samples in VTM were kept at 4°C after collection, and samples in NSLB were kept at room temperature until reaching the lab.

VTM samples were diluted in the lab in standard LB for inactivation of the virus and releasing of RNA into the medium. After 10-20 minutes in LB, RNA was extracted. NSLB samples were transferred directly to RNA extraction.

At the Sheba and Rambam Medical Centers, we extracted RNA by the Precision System Science MagLead 12gC with the MagDEA Dx (LV at Sheba and SV at Rambam). At Hadassah Medical Center we extracted RNA using three different commercial kits and platforms: MagNA Pure 96 kit (Roche Lifesciences) using Roche platform, Qiagen DSP virus/Pathogen kit using Qiasymphony platform, and MagDEA DX SSV kit (PSS, Japan) using the MagLead 12gC platform.

The Sheba and Hadassah RT-qPCR tests were conducted on the viral E gene, and the Rambam RT-qPCR was performed on the E, RdRp, and N genes (Seegene Allplex 2019-nCoV Assay). Human genes were also tested from the same sample: Sheba and Rambam tested the RNase P and Hadassah the ACTB. The limit of detection was Ct=40. For the calculation of Ct difference, a sample that was negative in one medium but its matched sample was positive, was defined as Ct=40.5. Each pair of samples from the same participant were always processed in the same 96 PCR plate.

All the raw data is available in the **Supplementary Material**.

## Results

### Using NSLB for sampling improves the sensitivity of SARS-COV-2 RT-qPCR tests

To compare the sensitivity of SARS-COV-2 tests in different collection media, we sampled patients once into VTM and then again into NSLB, conducted RT-qPCR on both samples, and compared the Cycle threshold (Ct) values of the matched samples from the same individual.

Overall, we tested 99 participants. Of them, 20 were healthy volunteers recruited as negative controls. All controls tested negative in both VTM and NSLB. The other 79 participants were COVID-19 patients (participants who previously tested positive at least once and were hospitalized or quarantined at the time of the study). Of these, 18/79 patients tested negative in both VTM and NSLB (of which 17 were from the recovering quarantined group). Those are likely patients who recovered and not false negatives since internal controls of human RNA were positive in all of these cases. One patient was negative for human RNA internal control (bad sampling) and was omitted from further analysis. The 60 patients who tested positive in either VTM, NSLB, or both, were further analyzed.

In 45/60 (75%) of the patients, the NSLB sample showed lower Ct value (higher viral titer) than its matched VTM sample, while in only 15/60 (25%) of patients, the Ct of VTM was lower (**Figure 1a-c)**. The average Ct difference in favor of NSLB was 2.1, CI [0.9,3.3] and this difference was statistically significant (t-test p-value = 8.4×10^−4^). 12/60 (20%) of the patients tested positive in NSLB but negative (below the limit of detection) in VTM, while only 6/60 (10%) tested positive in VTM but negative in NSLB. Overall, our results demonstrate a significantly higher sensitivity for sampling into NSLB over VTM. Importantly, NSLB was advantageous (lower Ct) over VTM in all the three different medical centers that participated in this study (each processed approximately a third of the samples, see **Methods**), although each had different teams of samplers and different lab protocols.

**Figure 1.**
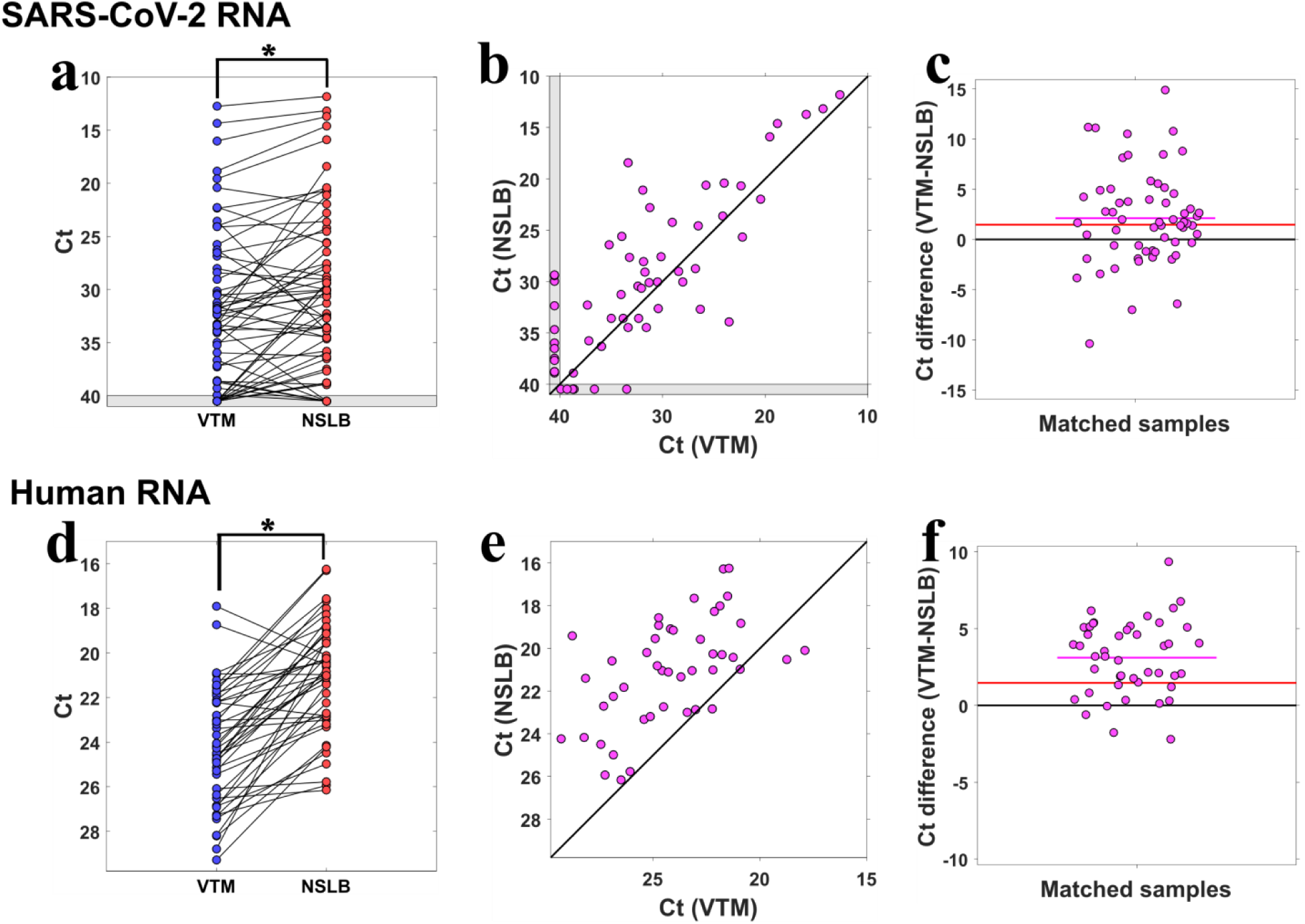
Comparison of sampling into VTM vs. NSLB. **(a)** Ct values of SARS-CoV-2 RNA in samples collected from patients into VTM (blue), linked by black lines to their NSLB matching samples from the same patients (red). The asterisk denotes a statistically significant advantage for NSLB. A grey zone adjacent to the axes marks results below the limit of detection. **(b)** Ct values of matched samples from the same patient in VTM (x-axis) vs. NSLB (y-axis). The diagonal black line represents theoretical equal performance of VTM and NSLB. **(c)** Ct difference (VTM-NSLB) for each sample pair from the same patient. The black line at y=0 represents theoretical equal performance of VTM and NSLB. The red line at y=1.5 represents equal performance of VTM and NSLB when controlling for VTM dilution. The magenta line shows the mean of differences. **(d,e,f)** Same as (a,b,c), but for human RNA.

When samples in VTM are processed in the lab they are typically inactivated in LB before entering the RNA extraction process. This inactivation dilutes the original sample, typically 2-3 fold. In our experiment, samples in VTM were diluted on average 2.8 fold in LB. Therefore, if NSLB is equal in performance with VTM with regards to biochemical properties alone, one would expect a baseline of log2(2.8)=1.5 lower Ct for NSLB. The observed average Ct difference of 2.1±0.6 (Mean±SE) was not significantly higher than 1.5 (t-test p-value = 0.28). This result suggests that at least most of the advantage of NSLB in our study was due to using a more concentrated sample for RNA extraction since no inactivation in LB (and hence dilution) was needed. We observed no significant difference between the two commercial NSLBs that we used (rank-sum test p-value = 0.52).

For 45 positive patients, we also performed RT-qPCR on human mRNA. NSLB samples showed on average 3.1 CI [2.4,3.8] lower Ct than their matched VTM samples (**Figure 1d-f**). Interestingly, this Ct difference was significantly higher than both zero (t-test p-value =5×10^−11^) and the 1.5 Ct expected difference when dilution is controlled for (t-test p-value =4×10^−5^). This suggests that the sampling into NSLB probably broke open more human cells over the time it took to transfer the samples to the lab (2-8 hours) compared to incubating VTM samples for 10-15 minutes in LB before RNA extraction. It also indicates that RNA in NSLB has not been degraded before reaching the lab.

### Samples in NSLB can be stored at room temperature at least 72 hours without a reduction in RT-qPCR performance

A possible concern when using NSLB for sampling is that RNA will be degraded prior to being processed in the lab, and thus the sensitivity of the test will decrease. To test the RNA stability over time, we repeated the RNA extraction of 24 samples from hospitalized patients (Sheba Medical Center) after 24 hours of storage at 4°C. Ct values were highly correlated for VTM samples between time 0h and time 24h (Pearson r=0.96), and also for NSLB samples (Pearson r=0.93), see **Figure 2a**. The advantage of NSLB over VTM did not significantly change after 24h (t-test P=0.63).

**Figure 2.**
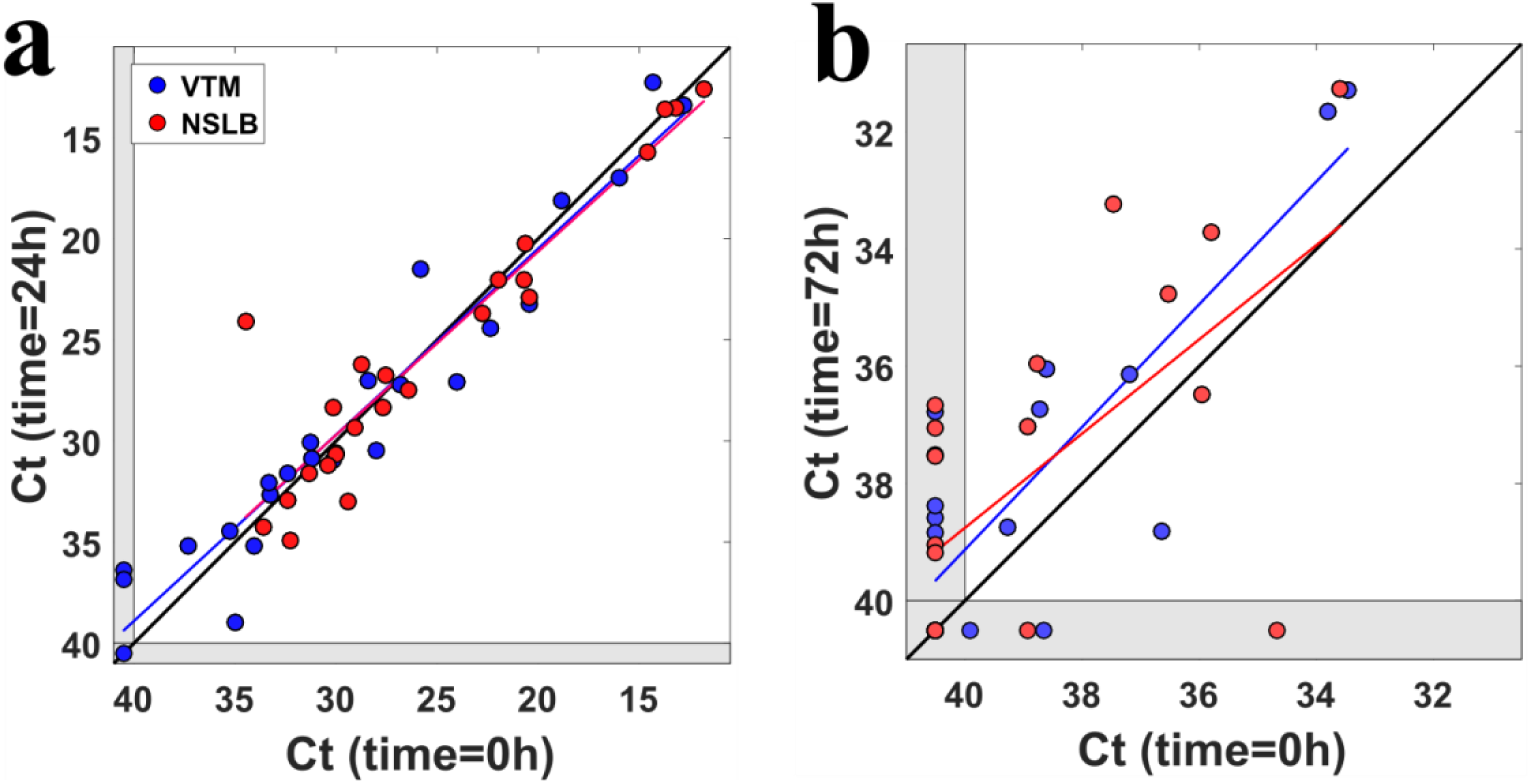
Detection of SARS-CoV-2 RNA at different times of sample storage. **(a)** Ct values at time=0h vs. time=24h for 24 hospitalized patients. Blue dots represent VTM samples and red dots represent NSLB samples. Colored lines are linear fits for VTM and NSLB. **(b)** Same as (a) but for 20 recovering quarantined patients, tested at times 0h and 72h.

For 20 other patients (recovering quarantined patients) we kept for 72 hours the NSLB samples at room temperature and the VTM samples at 4°C and then extracted RNA a second time and performed another RT-qPCR. Correlation of Ct values between time 0h and 72h were Pearson r=0.83 for VTM samples and Pearson r=0.63 for NSLB samples (**Figure 2b)**. Both VTM and NSLB samples showed a mean decrease of 1 Ct value after 72 hours (VTM: -1 CI [-1.6,-0.2], t-test p-value = 0.02, NSLB: -1 CI [-2.1,0.05], t-test p-value = 0.06), indicating that not only RNA was not degraded in either medium, but that perhaps even more detectable RNA was released into the mediums during these 72 hours. Also, the advantage of NSLB over VTM remained similar at 72h as it was at time 0h (t-test p-value=0.89).

### Sampling into NSLB eliminates the need to keep the swab in the tube

Sampling into NSLB is not sufficient for eliminating work under BSL2 conditions in the lab, since typically the swab is fully inserted into a patient’s mouth or nose but only the swab’s tip is dipped into the medium. In addition, keeping the swab in the tube poses a challenge for downstream automation procedures in the lab. Unlike VTM, the reagents in NSLB have the potential to release almost immediately most of the biological material from the swab into the medium. Therefore, we tested if biosafety and automation could be improved without impairing sensitivity, by discarding the swabs immediately upon sampling after a short swirl in NSLB. This method was described previously and proved to allow efficient detection of S. aureus [3], but has not been evaluated in the context of COVID-19. To test this, six patients were tested using two consecutive NSLB swabs collection, where one swab was retained in the tube, and the other was swirled-in the second tube and discarded. For RT-qPCR on the E gene of SARS-CoV-2, keeping the swab in showed on average only a slightly lower Ct value compared to the Ct of a parallel sample for which the swab was discarded (Mean of Ct differences = -0.27, CI [-1.7,1.2]). For RT-qPCR on the ERV3 human gene keeping the swab showed on average a higher Ct value compared to discarding the swab (Mean of Ct differences = 0.45, CI [-0.46,1.4]). For both viral and human genes the difference between keeping the swab and discarding it was not statistically significant (P=0.65, P=0.26 correspondingly).

## Discussion

Here, we showed that direct sampling into NSLB resulted in significantly higher sensitivity in SARS-CoV-2 RT-qPCR compared to sampling into VTM. We showed that samples can be kept in NSLB at room temperature for at least 72 hours prior to RNA extraction without significant loss of sensitivity. Moreover, we showed that swabs can be discarded at the sampling site after a few seconds of swirling in NSLB without a significant loss of sensitivity. We conducted the experiment in three different medical centers, using different personnel and lab protocols, and obtained similar results in all three centers, confirming that sampling into NSLB is robust to varying RNA extraction protocols.

The increased sensitivity was most likely gained by facilitating the introduction of a more concentrated sample into the RNA purification step. PCR tests for SARS-CoV-2 have shown a high variation of False Negative Rates (FNR), with some reports of up to 20%-30% FNR [4,5]. Increasing the sensitivity of PCR tests is especially important at the incubation stage, where a patient might not have developed yet a high viral load and might falsely test negative shortly before becoming infectious. In addition, pooling protocols that aim to increase the number of COVID-19 tests cause loss of sensitivity [6], and using NSLB could compensate for some of this sensitivity loss.

We showed the samples can be kept in NSLB at room temperature for at least 72 hours without degradation of RNA. This is important at times of high demand for COVID-19 testing and backlogs of samples waiting to be processed in labs. It can also simplify the transportation of large numbers of samples from remote areas, especially in countries where molecular diagnostic labs are far from the sampling sites.

We showed that when sampling into NSLB, it is possible to discard the swab at the sampling site without sacrificing the test’s sensitivity. This removes a major obstacle for automation of COVID-19 tests (as pipetting robots can be used without the hindrance of the swab in the tube). Since NSLB also circumvents the need to lyse the sample in the lab before RNA extraction, it opens up the possibility of sampling into tubes that can be inserted directly into an RNA extraction robotic pipeline. This can help to significantly increase the number and shorten the turn-around time of COVID-19 tests, an important measure to curb the COVID-19 pandemic since patients are most infectious prior and only shortly after symptoms onset [7].

Direct sampling into NSLB can reduce the risk of infecting the medical and lab personnel who conduct the sampling, transporting, and processing of the samples. In addition, the immediate inactivation of the virus in NSLB can improve the safety of a recently suggested swab pooling approach [8] that involves multiple sampling into one open tube. Since NSLB enables discarding of the swab after each sampling it can increase the swab pooling yield.

It should be noted, however, that there are several disadvantages to using NSLB. First, it is more expensive than VTM. Second, it typically contains reagents that require more careful handling than VTM in cases of spillover. Third, there are certain RNA purification protocols that are not suitable for samples in NSLB. For example, RNA extraction protocols that use bleach are not safe if used with NSLB that contains Guanidine. Lastly, NSLB does not allow for the culturing of the virus in the lab. However, with the current need for high throughput and rapid SARS-CoV-2 tests, and since most samples are not currently cultured, this disadvantage is negligible.

## Conclusion

We demonstrated that sampling of suspected COVID-19 patients directly into NSLB and then discarding the swab has significant advantages over using the current guideline with VTM. It increases the sensitivity of the test, increases safety, and facilitates better automatic handling of samples in the lab. These advantages can help to increase the number and rapidity of COVID-19 tests at a time it is most needed.

## Supporting information

Raw Data

## Data Availability

All the raw data is available in the Supplementary Material.

## Acknowledgments

We thank Prof. Laurence Freedman for statistical consultation.

## Conflicts of Interest

The authors declare no conflict of interest

## Notes

### Competing Interest Statement

The authors have declared no competing interest.

### Funding Statement

No external funding was received

### Author Declarations

IRBs of the Sheba, Hadassah, and Rambam Medical Centers in Israel have approved this research.

### Summary of Updates

List of authors corrected

